# Detection of Antibiotic-Resistant Bacteria, Resistance Determinants, and Mobile Elements in Surface Waters in Lebanon

**DOI:** 10.1101/2021.02.12.21251645

**Authors:** Jennifer Moussa, Edmond Abboud, Sima Tokajian

## Abstract

The prevalence of antibiotic-resistant bacteria in surface water in Lebanon is a growing concern and understanding the mechanisms of the spread of resistance determinants is essential. We aimed at studying the occurrence of resistant organisms and determinants in surface water sources in Lebanon and understanding their mobilization and transmission. Water samples were collected from five major rivers in Lebanon. 91 isolates were recovered out of which 25 were multidrug-resistant (MDR) and accordingly were further characterized. *Escherichia coli* and *Klebsiella pneumoniae* were the most commonly identified MDR isolates. Conjugation assays coupled with *in silico* plasmid analysis were performed and validated using PCR-based replicon typing (PBRT) to identify and confirm incompatibility groups and the localization of β-lactamase encoding genes. *E. coli* EC23 carried a *bla*_NDM-5_ gene on a conjugative, multireplicon plasmid, while *bla*_CTX-M-15_ and *bla*_TEM-1B_ were detected in the majority of the MDR isolates. Different ST types were identified including the highly virulent *E. coli* ST131. Our results showed a common occurrence of bacterial contaminants in surface water and an increase in the risk for the dissemination of resistance determinants exacerbated with the ongoing intensified population mobility in Lebanon and the widespread lack of wastewater treatment.

## Introduction

The rapid spread of antibiotic resistance is a worldwide concern. Previously, antimicrobial resistant bacteria were confined to hospitals and veterinary settings, but are now widely disseminated in aquatic environments including rivers (Koczura *et al*. 2012), sewage treatment plants (Ferreira Da Silva *et al*. 2007), hospital effluents (Spindler *et al*. 2012; Zhang *et al*. 2017), drinking (Walsh *et al*. 2011) and surface water (Pereira *et al*. 2013). Aquatic environments constantly receive pathogenic and potentially pathogenic bacteria from different sources including municipal, hospital, and agricultural waste and as such could be a reservoir for multi-drug resistant organisms (Egervärn *et al*. 2017; Sanganyado and Gwenzi 2019). The prevalence of antimicrobial resistant strains in such environments is a worldwide concern due to the potential health hazards to the people exposed to such aquatic environments through different activities (Baquero, Martínez and Cantón 2008).

Surface water is largely affected by natural processes, human activities and the fast growth of the population which deteriorates water quality and threatens its use (Wilbers *et al*. 2014). In Lebanon, surface water sources are used as the main supply for agricultural activities, electricity generation, leisure, and human consumption (Daou *et al*. 2018). However, most water sources in Lebanon are contaminated with raw sewage and industrial waste (Faour-Klingbeil *et al*. 2016).

Water supplies and sanitation have suffered a lot in Lebanon because of the civil war and later due to the influx of refugees living in informal settlements or under temporary living environments without access to safe water and sanitation (Daoud *et al*. 2018). In 1992, a study assessing water quality revealed that most of the water supply systems in Lebanon do not conform with the world health organization quality standards for community water supplies. In 2007, the study of coastal rivers showed high levels of fecal coliforms confirming a significant raw sewer water input (Houri and El Jeblawi 2007). Diab *et al*. (2018) further validated the contamination of different water sources in Lebanon including spring and well waters, which are directly consumed without treatment, and estuaries which are used in watering crops and animals. Diab *et al*. (2018) also studied the distribution of multi-drug resistant bacteria in water resources. Similarly, Tokajian *et al*. (2018), examined the impact of population influx on the prevalence of ESBL-producing *E. coli* recovered from river effluents in Lebanon, and revealed the prevalence of drug resistant organisms and introduction of new resistance patterns into water systems.

Reports revealing the role of environmental factors and the environment, water and sanitation, in propagating resistance determinants are still limited in Lebanon. The purpose of this study was to estimate the occurrence and determine the molecular characteristics of antimicrobial resistant organisms and resistance determinants with the ultimate goal being to understand mobilization and transmission in surface water and mitigate where possible the spread.

## Materials and Methods

### Study Design and Sample Collection

A total of 15 water samples were collected between 2017 and 2018 from Al Qa’a refugee camp and four other major river effluents across Lebanon with possible sewage contamination in the north, south, el beqaa, and Beirut (Figure 1). The collected volume was 5L using sterile containers, followed by 10x dilution before being inoculated on Blood and MacConkey agar. A total of 91 isolates were recovered and were named to reflect the type (genus and species) of the organism (EC= *E. coli*, EN= *Enterococcus* spp., KP= *Klebsiella pneumoniae*, SM= *Serratia marcescens*, SA= *Salmonella* spp., AB= *Acinetobacter baumanii*, DE= *Delftia* spp., HA= *Hafnia*, KL= *Kluyvera ascorbata*, SH= *Shewanella* spp., PR= *Providencia* spp., RA= *Raoultella* spp., CI= *Citrobacter* spp., EB= *Enterobacter* spp., AE= *Aeromonas* spp., PA= *Pseudomonas* spp.). Supplementary figure 5 shows a detailed description for all recovered isolates (Designation, location, date of isolation, and the type of the organism).

**Figure 1:**
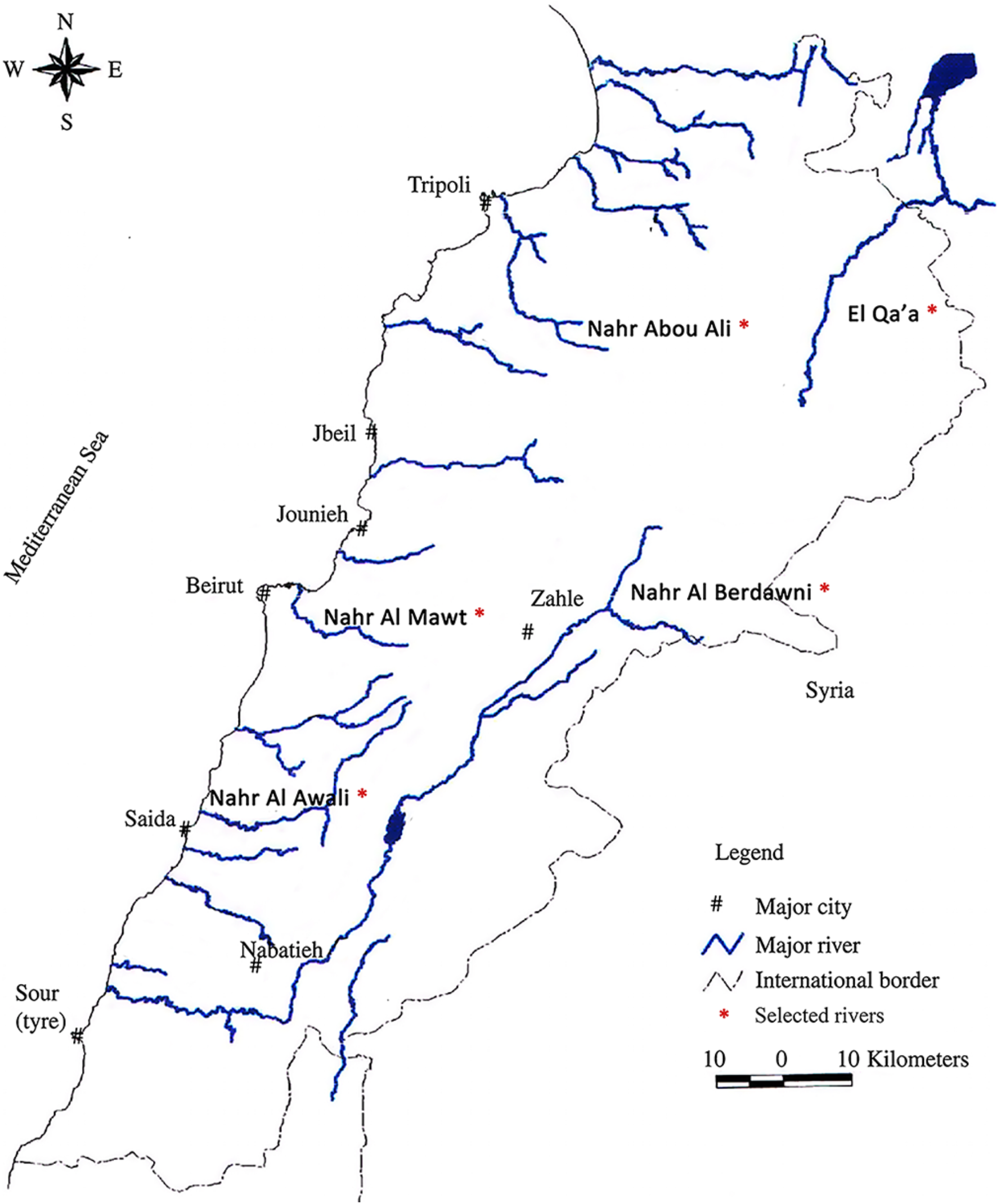
Geographical distribution of the chosen surface water sources included in this study; the red asterisk (*) indicates the rivers from which isolates were collected.

DNA extraction was performed using the NucleoSpin^®^ Tissue DNA extraction kit (Macherey-Nagel, Germany) following the manufacturer’s instructions and isolates were identified using 16S rRNA gene sequencing. *Escherichia coli* and *Klebsiella pneumoniae*, the two most commonly isolated organisms, were subjected to further characterization.

### Antimicrobial Susceptibility Testing

All Gram-negative isolates were tested for resistance using the disk diffusion assay on Mueller-Hinton agar using 29 different antimicrobial agents belonging to ten classes (Table 1). Gram-positive isolates were tested against 11 different antimicrobial agents (Table 2). Isolates identified as *Pseudomonas* spp. were tested against 13 antibiotics (data not shown). E-test strips (AB BIODISK, Solna, Sweden) were used with one recovered carbapenem-resistant *E. coli* (EC23) to determine the minimal inhibitory concentration (MIC) of ertapenem, imipenem and meropenem. All results were interpreted according to the Clinical Laboratory Standards Institute guidelines (CLSI 2018).

**Table 1:**
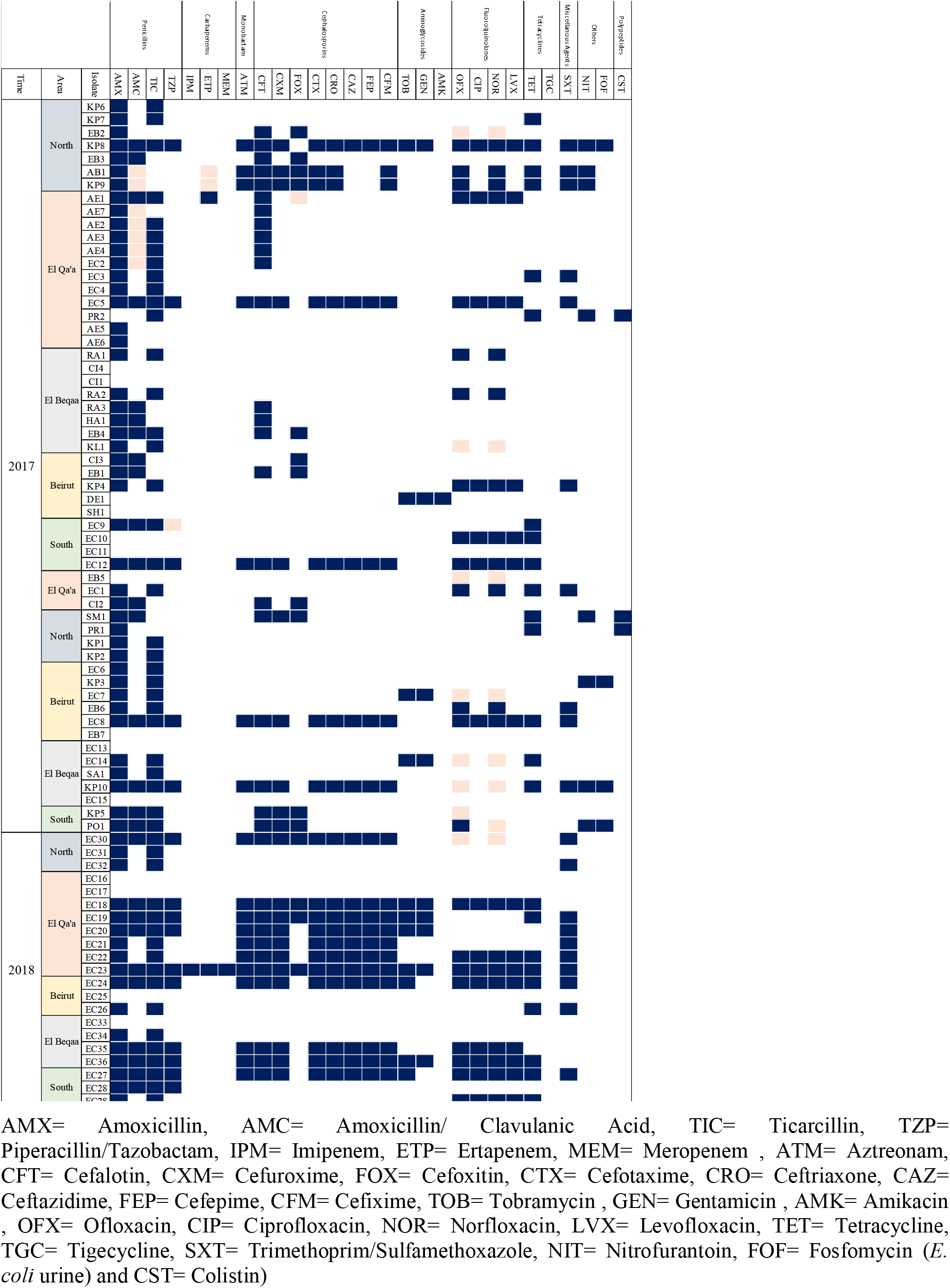
Antibiotic susceptibility results for all recovered Gram-negative organisms from the river effluents and El Qa’a refugee camp in Lebanon using 29 different antimicrobial agents covering ten different classes.

**Table 2:**
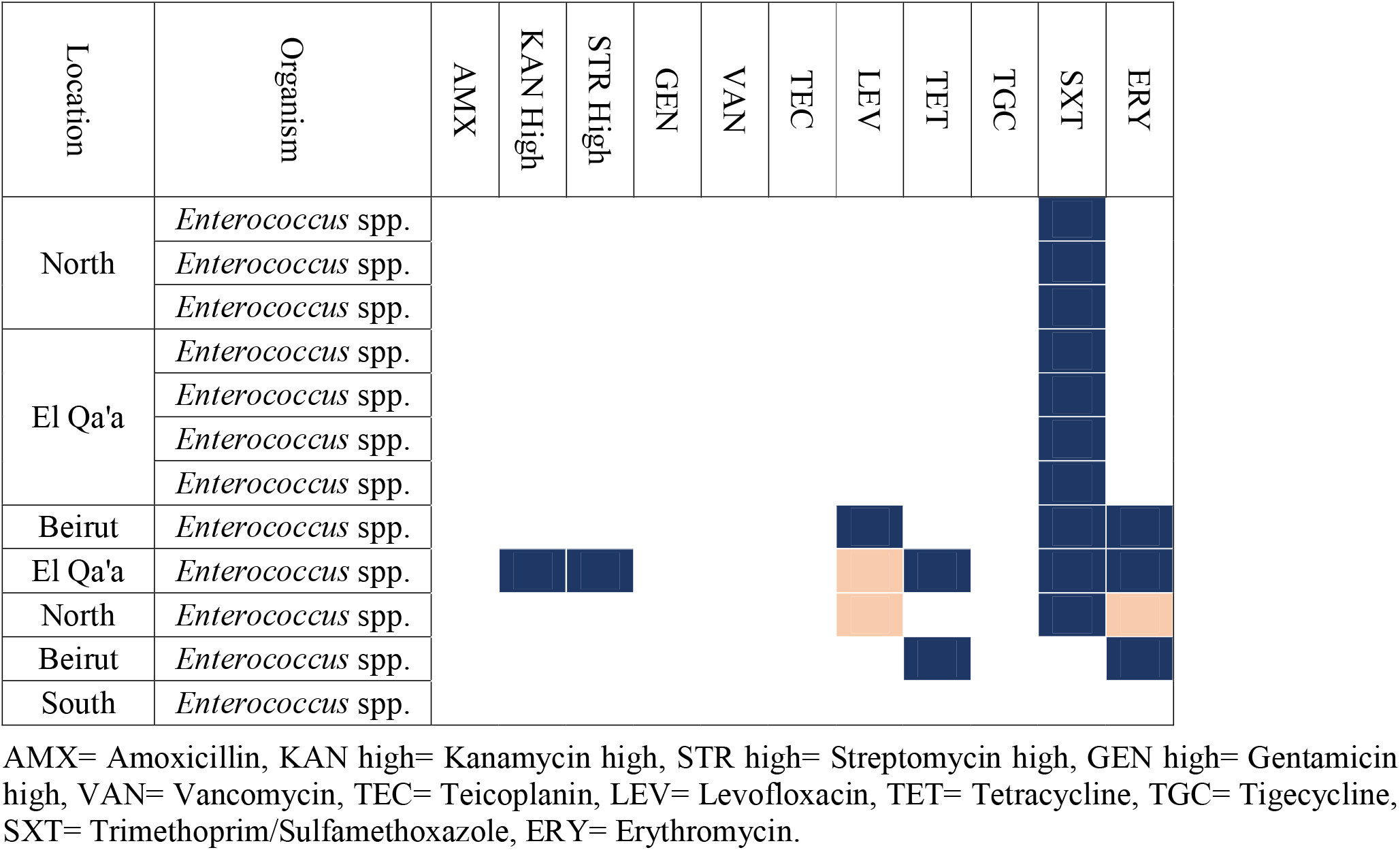
Antibiotic susceptibility results for all recovered Gram-positive organisms from the river effluents in Lebanon using 12 different antimicrobial agents covering eight different classes.

### Multi-locus sequence typing (MLST)

#### 1. K. pneumoniae

MLST was performed as described on the Institute Pasteur MLST database targeting seven housekeeping genes (*rpoB, gapA, mdh, pgi, phoE, infB*, and *tonB*) using primers with universal sequencing tails. Genes were sequenced using the universal oF and oR primer pair. STs were assigned using the Institute Pasteur database. (www.pasteur.fr/mlst).

#### 2. E. coli

The allelic profiles of the following seven housekeeping genes in *E. coli* were also determined: *adk, fumC, gyrB, icd, mdh, purA* and *recA* using primers with universal sequencing tails. Genes were sequenced using the same primers and STs were assigned using the MLST Warwick database (www.enterobase.warwick.ac.uk). The Achtman multilocus sequence typing (MLST) was also performed on all whole-genome sequenced isolates using MLST 2.0 database available on the Center for Genomic Epidemiology (CGE) (www.genomicepidemiology.org) (Larsen *et al*. 2012)

### PFGE fingerprinting

Genomic DNA plugs for *E. coli* and *K. pneumoniae* were prepared according to the standard PulseNet protocol (http://www.pulsenetinternational.org). Briefly, plugs were digested using *Xba*I (EC 3.1.24.4) (ThermoScientific, Waltham, MA, USA) for 2 h at 37°C. Electrophoresis was performed using the Bio-Rad laboratories CHEF DR-III system (Bio-Rad Laboratories, Bio-Rad Laboratories Inc., Hercules, CA, USA). *Salmonella enterica subsp. enterica serovar Braenderup* (ATCC® BAA664™) according to the standard PulseNet protocol (http://www.pulsenetinternational.org). Gels were stained with ethidium bromide. BioNumerics software version 7.6.1 (Applied Maths, St-Martens-Latem, Belgium) was used to analyze the PFGE profiles and pulsotypes were clustered through dice correlation coefficients with 1.5% optimization and 1.5% tolerance.

### Whole-genome sequencing

Based on the antimicrobial susceptibility test results, 25 isolates classified as being multi-drug resistant, acquired non-susceptibility to at least one agent in three or more antimicrobial categories (Magiorakos *et al*. 2012), or showed an interesting resistance pattern were chosen for in-depth molecular characterization through whole-genome sequencing. Library preparation was performed using the Illumina Nextera XT DNA Library preparation kit (Illumina, San Diego, CA, USA). Genomic DNA (gDNA) was used as input for library preparation. gDNA was subjected to end-repair, A-tailing, ligation of adaptors including sample-specific barcodes as per the manufacturer’s recommendation. Qubit 2.0 fluorometer (Invitrogen, Carlsbad, CA, USA) was used to quantify the resulting library which was sequenced on an Illumina MiSeq (Illumina, San Diego, CA, USA) with paired-end 500 cycles protocol to read a length of 250 bp. Genome assembly was performed *de novo* using SPAdes Genome Assembler Version 3.13.0 along with read error correction (Nurk *et al*. 2017). Quality control checks on the raw sequence data was performed using FastQC version 0.11.9 (Andrews 2010). The assembled draft genomes were annotated using RAST (http://rast.nmpdr.org/rast.cgi) (Aziz *et al*. 2008).

### Phylogenetic typing and Serotyping-*E. coli*

The phylogroup was determined as described previously (Clermont *et al*. 2013). The serotype was assigned using the SerotypeFinder 2.0. Results were further validated on GoSeqit (GoSeqit.com).

### PCR amplification and sequencing of *wzi* gene

To determine the capsular type, *wzi* gene typing was performed using wzi-TR and wzi-TF primers as previously described (Brisse *et al*. 2013). K-types were assigned using the Pasteur Institute database (http://bigsdb.pasteur.fr/klebsiella).

### Whole-genome based *in silico* typing and virulence and resistance profiling

*In silico* Plasmid and pMLST typing were performed on all whole-genome sequenced isolates using PlasmidFinder 1.3 and pMLST 2.0 by means of the Center for Genomic Epidemiology (CGE) online tools (Carattoli *et al*. 2014). Virulence genes were identified using VirulenceFinder 1.5 (Joensen *et al*. 2014). ResFinder v2.1 and the Comprehensive Antibiotic Resistance Database (CARD) were used to identify resistance genes (Zankari *et al*. 2012; Jia *et al*. 2017). Fimtyper 1.0 was used to identify the *E. coli* Fim type (Roer *et al*. 2017). Phage Search tool (PHASTER) was used for Phage identification (Arndt *et al*. 2016). IS-finder was used to identify insertion sequences (ISs) and IS-families (Siguier *et al*. 2006). plasmidSPAdes, version 3.12.0, was employed to generate separate plasmid contigs, and the output was visualized through Bandage and an assembly graph viewer. Mauve v2.4.0 was used for comparative genome alignment.

### Pan-genome and recombination analysis

To further study the diversity within *E. coli* and *K. pneumoniae*, genomes were annotated using Prokka (version 1.13) with a similarity cutoff e-value 10^−6^ and minimum contig size of 200□bp (Seemann 2014). Annotated GFF3 files were piped into Roary (version 3.12). Choosing a minimum BLASTp identity of 95 and core gene prevalence in all (>99%) of the isolates (Page *et al*. 2015)

A maximum-likelihood phylogenetic tree based on the core genome alignment was constructed using FastTree2. Gubbins version 2.2.1 was used to assess recombination events in core genes and to construct a maximum-likelihood tree using RAxML. One reference genome was included in each tree. *E. coli* str. K-12 subtr. MG1665 (accession # U00096.3) was used as a reference for the maximum-likelihood phylogenetic tree for all sequenced *E. coli* isolates, *Klebsiella pneumoniae* subsp. *pneumoniae* HS11286 (accession # NC_016845.1) was used as a reference for the *K. pneumoniae* isolates and *Escherichia coli* ST131 strain EC958 (accession # HG941718.1) was used when comparing *E. coli* isolates belonging to ST131. The resulting phylogenetic trees, isolates metadata along with the pan-genome fingerprints of the isolates, core genome SNPs, and recombination hotspots were visualized using Phandango V1.3.0.

### Plasmid typing and Conjugation assay

Plasmid incompatibility groups were identified by the PCR-based replicon-typing method using the DIATHEVA PCR-Based Replicon Typing (PBRT) kit (Diatheva, Fano, Italy).

### Conjugation Assays

#### Conjugation-*bla*_NDM-5_

Conjugal transfer of plasmid-borne *bla*_NDM-5_ from *E. coli* EC23 isolated from El Qa’a refugee camp was assessed by broth culture mating assay using *E. coli* J53 as a recipient as previously described by Inoue, Itoh and Mitsuhashi (1983) with some modifications. Exponential phase growing donor and recipient strains were mixed at a volume ratio of 1:3. The mating mixture was incubated for 2 h at 37°C and then plated on MacConkey agar supplemented with 100 μg mL^-1^ sodium azide and 10 μg mL^-1^ meropenem. Plasmid DNA was extracted from the transconjugant using the plasmid Mini Kit (Qiagen GmbH, Hilden, Germany). PBRT and PCR for the *bla*_NDM-5_ were performed on the plasmid DNA

#### Conjugation-*bla*_CTX-M-15_

Twelve isolates from different dates and locations were shown to carry *bla*_CTX-M-15_ (Figure 2). Conjugation experiments were conducted by the broth mating method using *bla*_CTX-M-15_ containing strains as donor and sodium azide-resistant *E. coli* J53 as the recipient, as previously described (Inoue, Itoh and Mitsuhashi 1983). The mating mixture was incubated for 2 h at 37°C and MacConkey agar supplemented with sodium azide (100 μg mL^- 1^) and cefixime (5 μg mL^-1^) was used to select for transconjugants. Plasmids were typed according to their incompatibility group using PBRT (Carattoli 2009).

**Figure 2:**
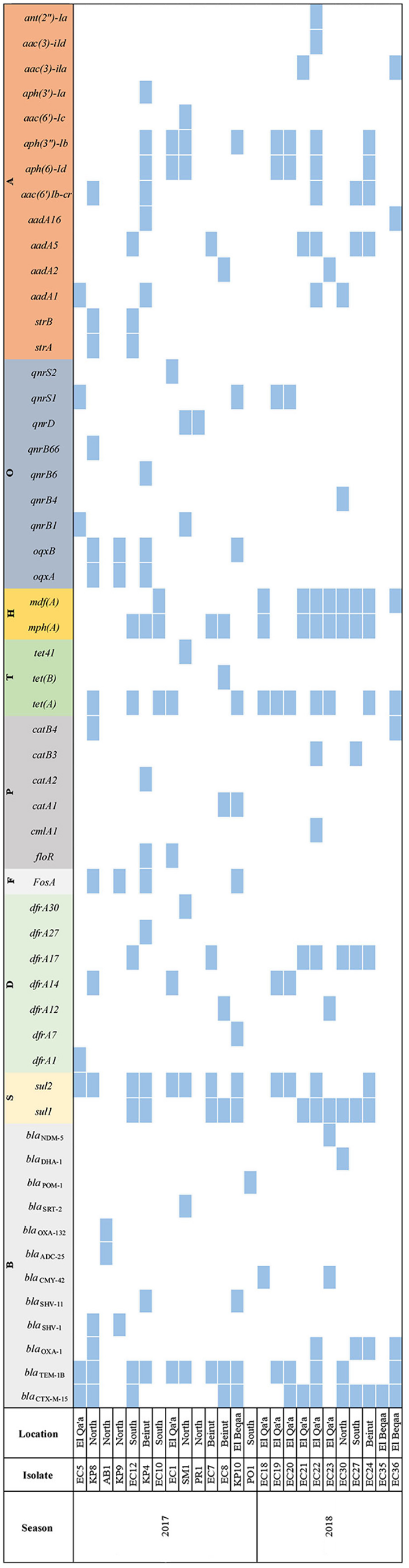
Types and distribution of resistance determinants as assigned using ResFinder v1.2 The used antimicrobial categories were labeled as follows: B, β-lactam resistance genes; S, sulfonamide resistance genes; D, trimethoprim resistance genes; F, fosfomycin resistance genes; P, chloramphenicol resistance genes, T, tetracycline resistance genes; H, macrolide resistance genes; O, quinolone resistance genes and A, aminoglycoside resistance genes.

Additionally, a conjugation assay was conducted using *E. coli* EC7 as a recipient and *E. coli* EC36 as a donor isolate. Selection on trimethoprim (50 μg mL^-1^) and cefixime (5 μg mL^- 1^) for *bla*_CTX-M-15_ positive transconjugants was used.

## Results

### Bacterial identification

A total of 91 organisms were isolated between 2017 and 2018 from four major sites: North (19/91, 20.9%), South (10/91, 10.9%), El Beqaa (17/91, 18.7%) and Beirut (17/91, 18.7%), in addition to El Qa’a refugee camp (28/91, 30.8%). Among the recovered isolates, 36/91 were identified as *E. coli* (39.6%), 12/91 *Enterococcus* spp. (13.2%), 11/91 *K. pneumoniae* (12.1%), the remaining 32/91 (35.1%) belonged to 13 different genera (Supplementary figure 1).

### Antibiotic Resistance Profiles

Most of the recovered isolates (86.8%; n= 79/91) were Gram-negative. *Pseudomonas aeruginosa* was tested using a different panel of antimicrobial agents and as such data obtained was analyzed apart from the other recovered Gram-negative isolates. The disk diffusion assay revealed that 81% (62/77) of the isolates were resistant to amoxicillin, 64% (49/77) to ticarcillin, and 44% (34/77) to cefalotin. All isolates were sensitive to tigecycline. *E. coli* EC23 isolated from El Qa’a refugee camp in 2018 was the only isolate resistant to ertapenem, meropenem, and imipenem (Table 1)

In the Gram-positive isolates however, the majority (83.3%; n= 10/12) were resistant to trimethoprim/sulfamethoxazole, while 25% (3/12) showed resistance to erythromycin. All the isolates were sensitive to amoxicillin, vancomycin, teicoplanin and tigecycline (Table 2).

Twenty-five isolates, mainly *E. coli* (17/25) and *K. pneumoniae* (4/25), were identified as being MDR, showed resistance to at least one agent in three or more antimicrobial categories (Magiorakos *et al*. 2012), and subsequently were chosen for further in-depth molecular characterization using whole-genome sequencing (Table 3). In addition, two colistin resistant isolates were detected and they were identified as non-pigmented *Serratia marcescens* and one *Providencia alcalifacians* which are intrinsically resistant to this antibiotic.

**Table 3:**
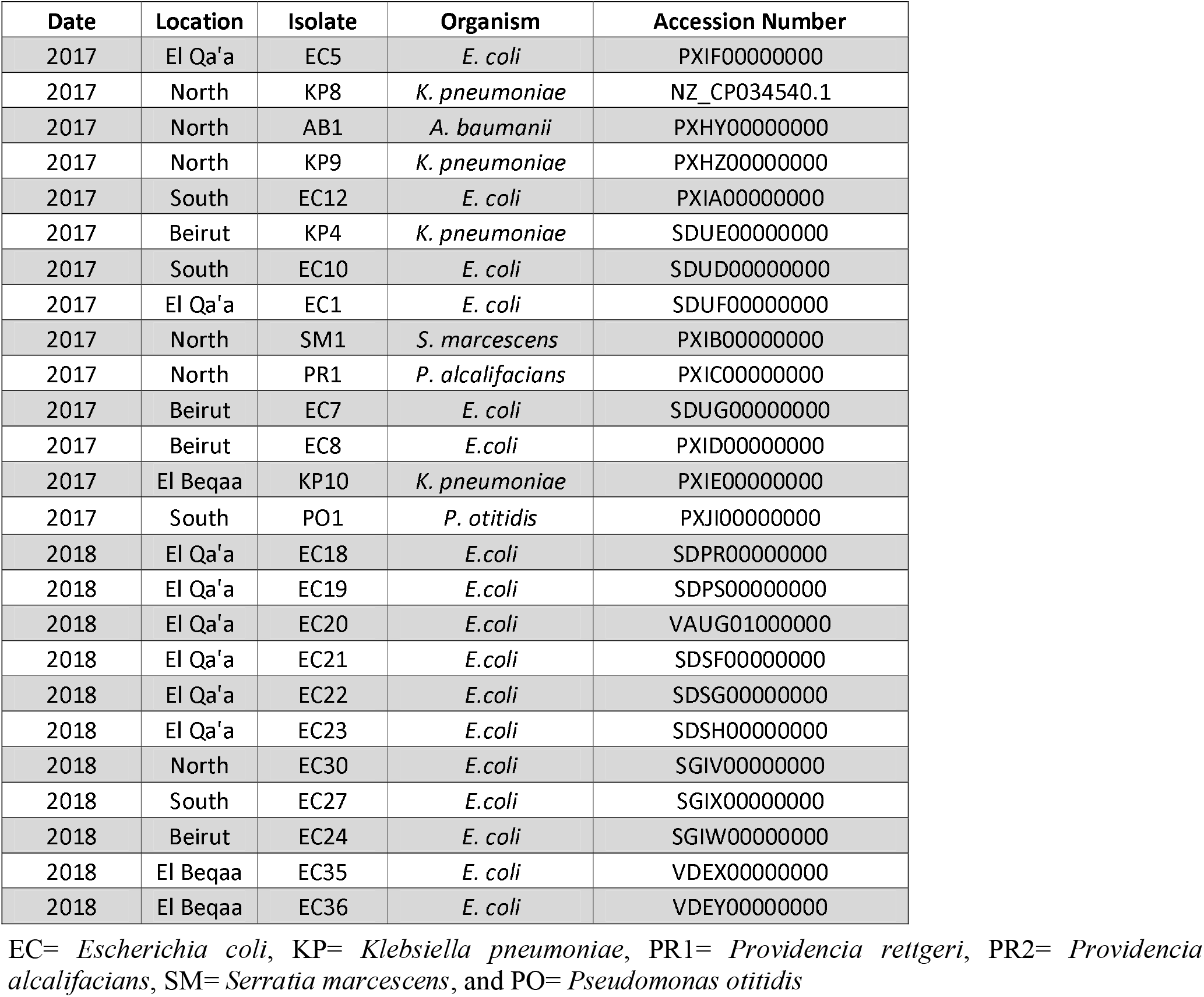
Detailed overview on the whole genome sequenced isolates that we chose for further characterization including the: season, year, site of isolation and NCBI accession numbers.

### Antibiotic Resistance Determinants

The antibiotic susceptibility testing results were further validated *in silico* through the detection of resistance determinants. A total of 56 genes conferring resistance to one of the nine tested categories of antimicrobial agents (β-lactams, fluoroquinolones, fosfomycin, sulphonamides, trimethoprim, tetracycline, phenicol, macrolides and aminoglycosides) was detected.

We found 12 different variants of genes linked to β-lactam resistance. The *bla*_TEM-1B_ gene was the most prevalent within the MDR isolates, and was detected in 14 isolates (10 *E. coli*, 3 *K. pneumoniae*, and 1 *S. marcescens*). The *bla*_CTX-M-15_ found in 13 isolates (46%), out of which five were additionally positive for *bla*_OXA-1_. Only one gene conferring resistance to carbapenems, namely *bla*_NDM-5_ was detected in this study from *E. coli* EC23. Fourteen different aminoglycoside resistance genes were also found. The most common were *aph(3”)-lb* (8/25; 32%) and *aph(6)-Id*. We additionally, detected phenicol, macrolide, tetracycline, sulphonamide, fosfomycin and quinolone resistance determinants (Figure 2).

### Plasmid typing

The combined results from PBRT and *in silico* replicon typing using PlasmidFinder 1.3 revealed 15 different replicons with the ones most prevalently detected Inc groups being IncFII (22.39%, 15/67), IncFIA (19.4%, 13/67), IncFIB (16.42%, 11/67) and IncFIB(K) (8.96%, 6/67). At least two different Inc groups were identified in the majority of the tested isolates (18/25, 72%). The highest number of Inc groups (n=7) was detected in EC18 isolated from El Qa’a refugee camp. The plasmidSPAdes version 3.12.0 was used to reconstruct the plasmids from paired-end reads. Based on that we showed the presence of a large multi-replicon plasmid having IncFIA, IncFIB, IncFII and InI1γ and carrying *bla*_CMY-42_ and *tet*A resistance determinants. Another smaller plasmid, missed due to Illumina short read sequencing and/or miss-assembly, carrying IncY. IncU, and IncI1α replicons were also detected.

### Molecular Typing

*E. coli* and *K. pneumoniae* were typed using PFGE, MLST, phylogrouping and serotyping to study genetic relatedness. The Achtman method (https://enterobase.warwick.ac.uk/species/ecoli) was used to determine the MLST for *E. coli*, based on which we found that ST131 (16.7%, 6/36) and ST69 (11.1%, 4/36) were the most common. We also identified 17 other STs including ST405, ST394, and ST410. Four of the isolates did not match any allelic profile and as such were assigned new STs. Using Clermont *et al*. (2013) classification scheme, we were able to phylotype the 36 *E. coli* isolates and accordingly were distributed into five phylogroups: A (27.8%; n=10/36), B2 (36%; n=13/36), D (17%; n=6/36), C (11.1%; n=4/36), and E (5.6%; n=2/36) (Figure 3).

**Figure 3:**
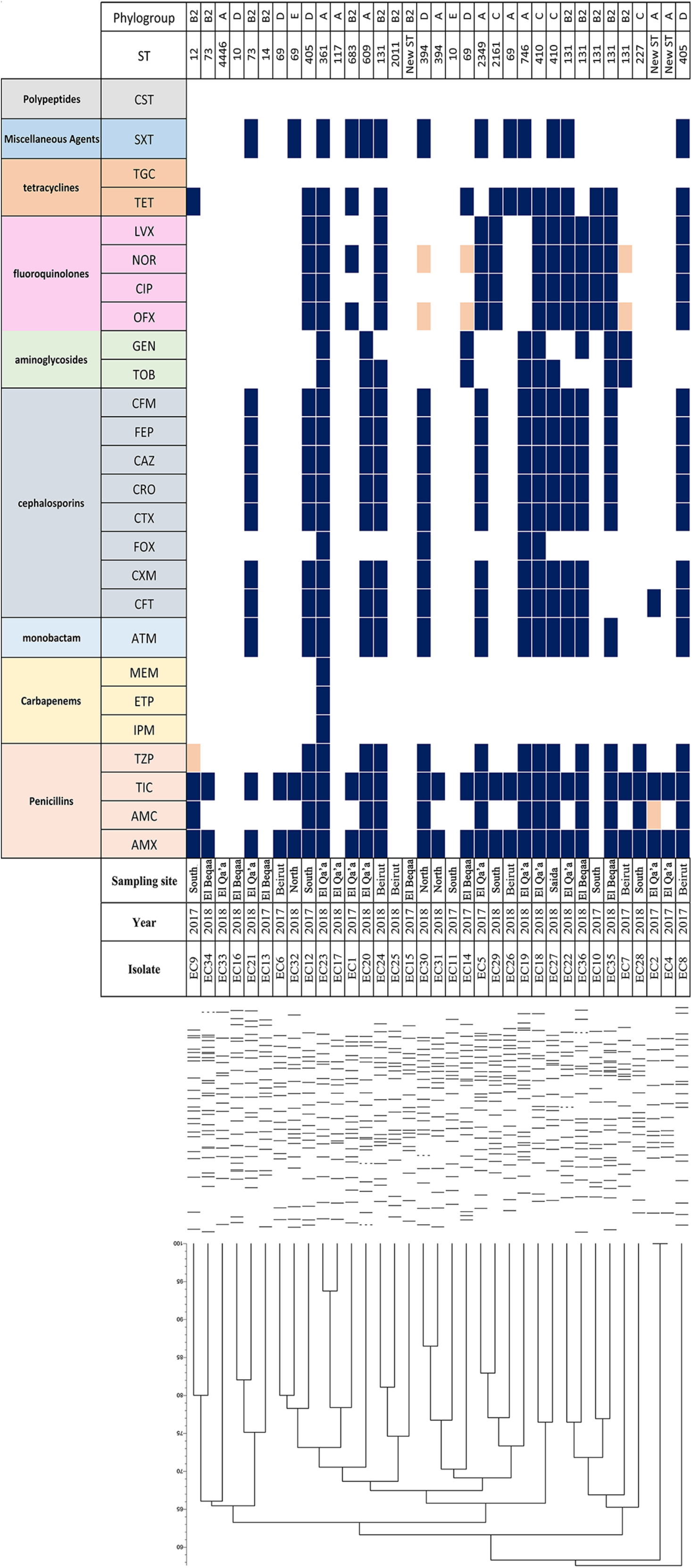
Detailed characterization of all *E. coli* isolates. The dendrogram was generated using Bionumerics software 7.6.2 showing the relatedness of the isolates based on their banding patterns generated by XbaI restriction digestion. Dark blue= Resistant, white= sensitive and light orange= intermediate resistant. AMX= Amoxicillin, AMC= Amoxicillin/ Clavulanic Acid, TIC= Ticarcillin, TZP= Piperacillin/Tazobactam, IPM= Imipenem, ETP= Ertapenem, MEM= Meropenem, ATM= Aztreonam, CFT= Cefalotin, CXM= Cefuroxime, FOX= Cefoxitin, CTX= Cefotaxime, CRO= Ceftriaxone, CAZ= Ceftazidime, FEP= Cefepime, CFM= Cefixime, TOB= Tobramycin, GEN= Gentamicin, OFX= Ofloxacin, CIP= Ciprofloxacin, NOR= Norfloxacin, LVX= Levofloxacin, TET= Tetracycline, TGC= Tigecycline, SXT= Trimethoprim/Sulfamethoxazole and CST= Colistin, EC= *E. coli*, ST= sequence type.

Serotypes of sequenced *E. coli* isolates were also determined *in silico* using SerotypeFinder 2.0. ST131 isolates (EC10, EC22, EC24, EC35, and EC36) belonged to O25b:H4 serotype except for EC7 which was typed as O16:H5b. Other serotypes were also found including: O102:H6 (ST405; EC8 and EC12). EC5 (ST2349), EC19 (ST746), and EC27 (ST10) however, had untypeable O type gene.

Using PFGE, a total of 34 pulsotypes were obtained. Isolates with the same ST did not cluster together; EC24, for example, typed as ST131 clustered away from the other five isolates from the same ST (Figure 3). It is noteworthy that EC3 (El Qa’a refugee camp) was untypeable.

With *K. pneumoniae*, seven different STs and K-types were identified while some were untypable. KP9 and KP6 belonged to ST76 and were linked to K-type 31. KP9 and KP6 additionally, shared a 96% similarity despite the difference observed in their susceptibility patterns. (Figure 4).

**Figure 4:**
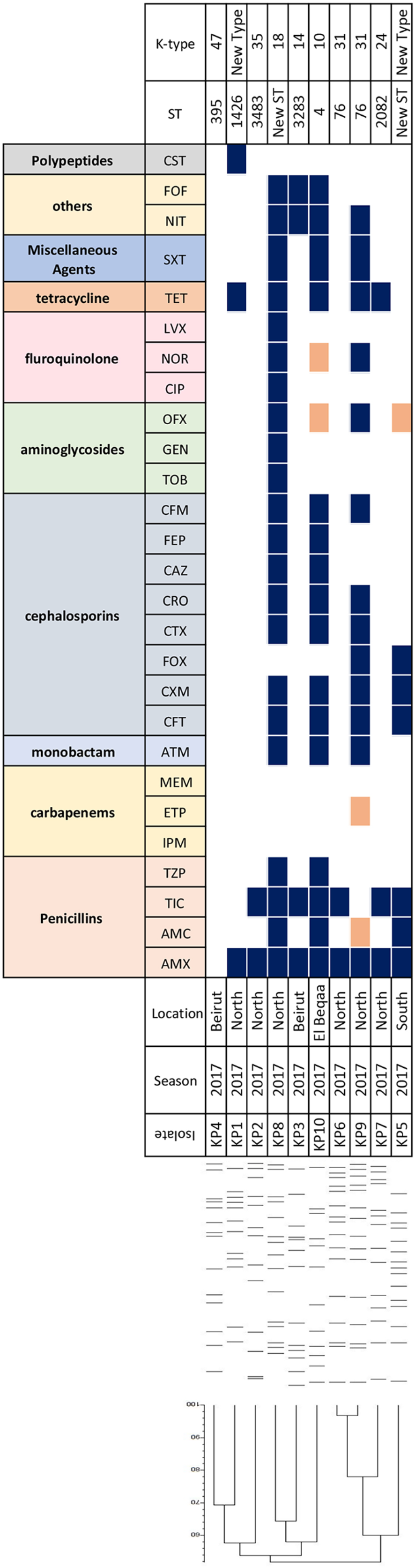
Detailed characterization of all *K. pneumoniae* isolates. The dendrogram was generated using Bionumerics software 7.6.2 showing the relatedness of the isolates based on their banding patterns generated by XbaI restriction digestion. Dark blue= Resistant, white= sensitive and light orange= intermediate resistant. AMX= Amoxicillin, AMC= Amoxicillin/ Clavulanic Acid, TIC= Ticarcillin, TZP= Piperacillin/Tazobactam, IPM= Imipenem, ETP= Ertapenem, MEM= Meropenem, ATM= Aztreonam, CFT= Cefalotin, CXM= Cefuroxime, FOX= Cefoxitin, CTX= Cefotaxime, CRO= Ceftriaxone, CAZ= Ceftazidime, FEP= Cefepime, CFM= Cefixime, TOB= Tobramycin, GEN= Gentamicin, OFX= Ofloxacin, CIP= Ciprofloxacin, NOR= Norfloxacin, LVX= Levofloxacin, TET= Tetracycline, TGC= Tigecycline, SXT= Trimethoprim/Sulfamethoxazole and CST= Colistin, EC= *E. coli*, ST= sequence type.

### Genome Statistics and Comparative Analysis

Pan-genome analysis of all the MDR *E. coli* isolates revealed the presence of a total of 10,315 unique protein coding sequences. Genes identified as part of the core genome were detected in all and constituted 3,154 protein coding sequences. Genes present in 15-95% of isolates were designated as being part of the shell genes and constituted 2,451 protein coding sequences, while those present in <15% constituted 4,710 protein coding sequences. Maximum-likelihood phylogenetic tree constructed based on single nucleotide polymorphisms (SNPs) placed the isolates in three different clusters. WgSNPs-based phylogenetic typing however, clustered the isolates based on differences in their STs, phylogroups, serotypes, and FimH. Again, three clusters were observed with one having all the isolates belonging to phylogroup B2 which included six isolates typed as ST131 and another as ST73. Isolates belonging to phylogroup D (ST405 and ST394), A, and, C also clustered together (Supplementary figure 2).

We also constructed a SNP-based maximum-likelihood phylogenetic tree for the isolates typed as ST131. We used a reference genome of the same MLST type, and of phylogroup B2 and serotype O25b:H4 (Accession # HG941718.1). EC7 with serotype O16:H5 clustered separately from the other isolates (Supplementary figure 3).

Using BLASTn search against the non-redundant nucleotide database we were able to associate the highly recombinant region to the *kpsFEDUCS* region 1 operon in *E. coli* ST131 isolates.

### Conjugation assays

All CTX-M-15 bearing isolates were tested for transferability of the plasmid carrying the *bla*_CTX-M-15_ gene using *E. coli* J53. Transconjugants containing CTX-M-15 encoding plasmids were extracted from five isolates. Among the five transferable CTX-M-15 producers, the most prevalent Inc type was IncF (n=3) with only one *bla*_CTX-M-15_ being carried on IncI1⍰ plasmid, and another on IncB/O/K/Z plasmid. IncF plasmids were also positive for other β-lactamase encoding genes such ad *bla*_TEM-1B_, *bla*_CMY-42_, *bla*_DHA-1_, and *bla*_OXA-1._ *bla*_NDM-5_ on the other hand, was detected on a 120 kb plasmid and upon conjugation *E. coli* J53 showed resistance to meropenem, imipenem, and ertapenem.

*bla*_CTX-M-15_ genes were also chromosomal in almost half of the isolates (51.3%) with two (EC30 and EC35) having it on SSU5_Salmonella phage (Supplementary figure 4, Table 4). In addition, the *in silico* analysis revealed that *bla*_ADC-1_ and *bla*_OXA-132_ in *Acinetobacter baumannii*, and *bla*_SRT-2_ and *bla*_TEM-1B_ in *S. marcescens* were all chromosomal (**Error!**

**Table 4:**
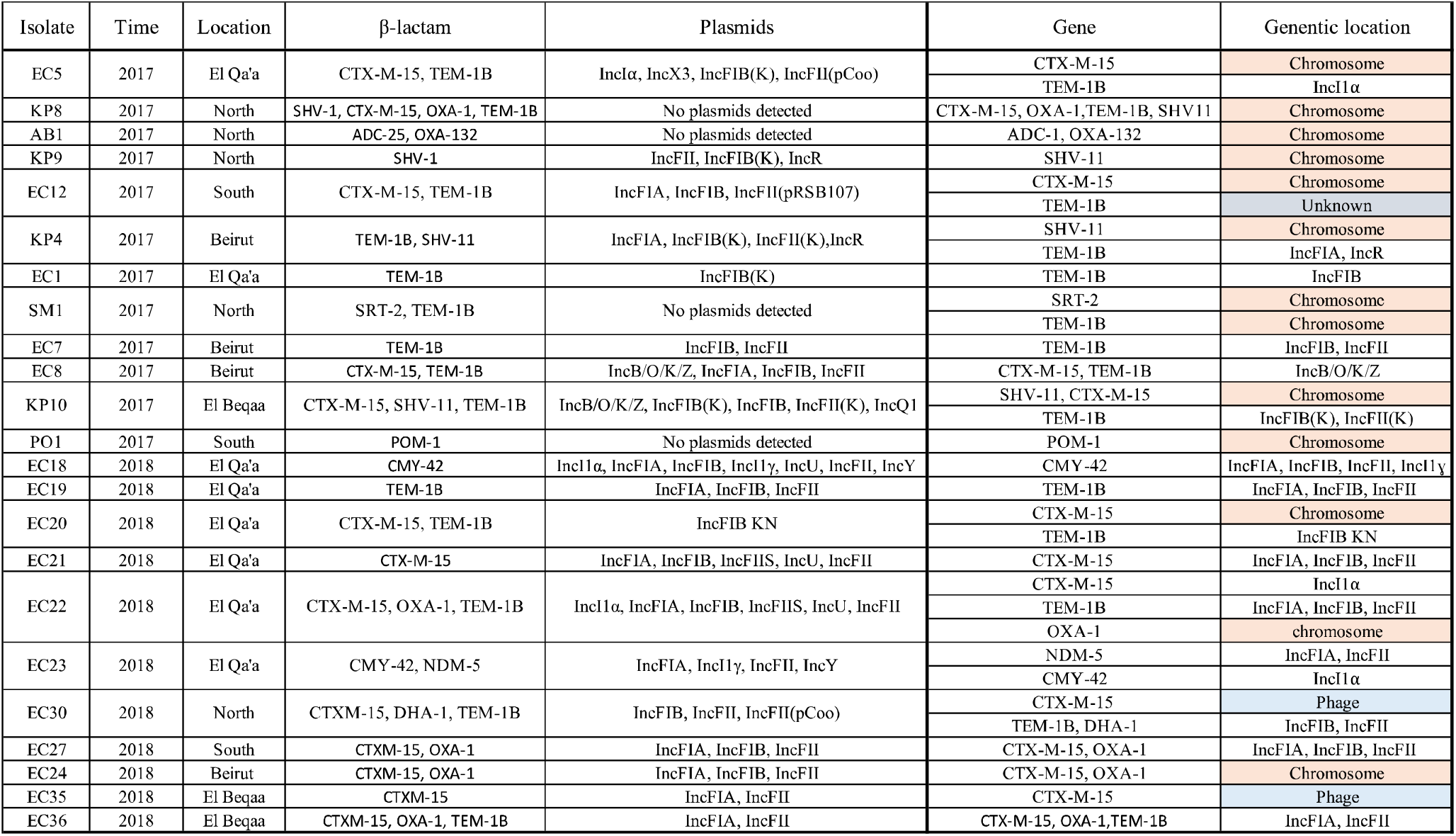
Distribution of β-lactamase encoding genes, plasmid typing results and gene locations. *in silico* analysis was done using ResFinder v1.2, while plasmid Inc group determination was through PBRT-based PCR assays combined with *in silico* analysis using PlasmidFinder 1.3. Gene location was determined using conjugation assay and *in silico* analysis.

### Reference source not found.)

We also assessed the mobility of the *bla*_CTX-M-15_ by performing a conjugation assay using EC36 as donor and to EC7 as recipient. The plasmid carrying *bla*_CTX-M-15_ was mobilized through the performed conjugation assay as also shown by the detected cefixime resistance in EC7. PBRT and PCR assays also revealed that EC7 was positive for the *bla*_CTX- M-15_ and had three replicons (IncFIA, IncFIB and IncFII).

## Discussion

Antibiotic resistance is a worldwide concern and the spread of drug-resistant bacteria through different environments including water may constitute a route for the dissemination of drug-resistant pathogens (Egervärn *et al*. 2017). The prevalence of antibiotic-resistant bacteria in surface water in Lebanon is a growing concern with the lack of collection and domestic wastewater management strategies. According to the Central Administration of Statistics, only 37% of the buildings in Lebanon are connected to a sewer networks, the rest either use cesspools or septic tanks or release raw sewage directly into the environment (Ministry of Environment, ECODIT). Community and hospital effluents could contaminate surface water and increase the microbial load (Houri and El Jeblawi 2007). In this study, we aimed at determining the role of surface water in the mobilization of resistance determinants. Our results showed a common occurrence of drug-resistant organisms in surface water including ESBL- and carbapenemase-producing *Enterobacteriaceae* being exacerbated by population mobility and the absence of wastewater treatment. Sewage contaminated surface water samples from river effluents were collected from across Lebanon. Antibiotic susceptibility testing was performed and MDR isolates (Magiorakos *et al*. 2012), or the ones that showed an interesting resistance profile, were further characterized using whole-genome sequencing. Ninety-one isolates in total were recovered and 25 were whole-genome sequenced and further characterized.

*E. coli* (39%; n= 36/91) was the most commonly recovered organism. This finding was in accordance with a previous study by Tokajian *et al*. (2018), assessing bacterial loads from sewage contaminated surface water samples from Lebanon. The second most prevalent was *Enterococcus* spp. (13%; n= 12/91) despite the fact that in general they are not frequently recovered from water (Cabral 2010). Their isolation from aquatic environments was linked to livestock, poultry, hospital, and municipal sewage, and wildlife fecal contamination (Byappanahalli *et al*. 2012).

*S. marcescens* recovered in this study was a non-pigmented biotype and positive for TEM-1B and SRT-2. It’s noteworthy that both pigmented and not-pigmented biotypes were previously detected with the non-pigmented being more virulent (Roy, Ahmed and Grover 2014). Hospital outbreaks were also linked to the non-pigmented biotype and which was in contrast to the environmental strains that produced a red pigment, prodigiosin (Kurz *et al*. 2003).

MLST typing, revealed that *E. coli* recovered in this study belonged to 20 unique ST types. *E. coli* typed as ST131, one of the high-risk clones associated with community onset infections (Mathers, Peirano and Pitout 2015), was the most common, and was detected in five out of the six collection sites. ST131 was also previously recovered from dug and spring wells and estuaries from Lebanon as well as different water sources worldwide (Bréchet *et al*. 2014; Chen *et al*. 2016; Egervärn *et al*. 2017). Interestingly, clinical and ecological studies have shown that CTX-M-15 producing ST131 could be rarely recovered from the environmental or veterinary settings (Pitout and DeVinney 2017). Add to it the fact that ST131 was frequently recovered from clinical settings in Lebanon, and which could further suggest contamination linked to human-activities.

Moreover, the five ST131 isolates were of serotype O25b:H4 and of the FimH30 lineage. FimH30 is the most prevalent lineage within ST131; it is named as such because it contains the H30 variant of the type 1 fimbrial adhesin gene (Mathers, Peirano and Pitout 2015). FimH30 also includes around 70% of the recent fluoroquinolone resistant ST131 *E. coli* isolates and which were implicated in the global dissemination of the fluoroquinolone resistant ST131 associated with extra intestinal infections (Johnson *et al*. 2010). In contrast, only one of the recovered isolates in this study (EC7) was of a different serotype namely O16:H5 and which belonged to the FimH41 lineage. A lineage that encompasses only 1-5% of the ST131 *E. coli* (Johnson *et al*. 2014).

Phylogrouping showed that B2 (36%) was the most common followed by phylogroup A (27.8%), and D (17%). Commensal *E. coli* isolates generally belonged to phylogroups A and B1, while the extra intestinal ones which were more pathogenic to B2 and D (Johnson *et al*. 2001). Phylogroups A and B1 however, were more frequently recovered from aquatic environments than phylogroups B2 and D (Figueira, Serra and Manaia 2011).

Amongst the whole-genome sequenced isolates in this study, 80% showed resistance to ticarcillin and 72% to third-generation cephalosporins and aztreonam. *bla*_CTX-M-15_ was the most common detected ESBL in both *E. coli* and *K. pneumoniae. bla*_CTX-M-15_ prevalence confirmed previous results (Diab *et al*. 2018; Tokajian *et al*. 2018).

The frequent detection of ESBL in clinical settings and the environment could reveal mobilization through horizontal gene transfer. Such transmission is favored by mobile genetic elements, specifically plasmids. IncF plasmids have a broad host range facilitating mobility and spread of resistance determinants (Villa *et al*. 2010; Zhao and Hu 2013). IncF plasmids were implicated in the spread of TEM, OXA, CMY, and KPC and often conferring multi-drug resistance (Villa *et al*. 2010; Carattoli 2011).

In addition, one NDM-5 positive *E. coli* (EC23, ST361) was recovered from El Qa’a refugee camp. NDM-5 was first described in a clinical *E. coli* isolate in 2011 from the United Kingdom (Hornsey, Phee and Wareham 2011). It was also detected in Montpellier, France from an urban river water sample (Almakki *et al*. 2017) and in India from a hospital-linked sewage water. In Lebanon however, *bla*_NDM-5_ was only linked so far to *K. pneumoniae* with clinical origin (Parvez and Khan 2017; Nawfal Dagher *et al*. 2019). *bla*_NDM-5_ in this study was on a conjugative multi-replicon plasmid (IncFIA, IncFII). *bla*_NDM-5_ commonly was found associated with IncX3 (Almakki *et al*. 2017), but was also linked to IncF plasmids from environmental samples in Myanmar (Sugawara *et al*. 2019) and from wastewater treatment plant effluent in Switzerland (Zurfluh *et al*. 2018).

The detection of MDR *E. coli* in surface water along with mobile element carrying resistance determinants highlights and confirms the role of water and lack of proper sanitation in the propagation and mobilization of resistance determinants. Our results suggested and confirmed that surface water could act as an important reservoir facilitating the spread of resistance determinants. We have also characterized the genetic basis of antibiotic resistance of ESBLs as well as other antimicrobial agents. Efforts in the future should be focused at building waste treatment plants, minimize pollution from agricultural sources, and control and mitigate human activities linked to introducing antibiotic resistance into the environment, which is an urgent, challenging, and pressing task.

## Data Availability

Included in manuscript and submitted documents

## Funding

This work was supported partially by the School of Arts of Sciences Research and Development Council at the Lebanese American University and by the National Council for Scientific Research (Grant #756).

## Conflict of Interest

None declared

## Acknowledgments

None.

## Supplemental Files

**Supplementary figure 1:** Distribution of organisms isolated from river effluents (El Beqaa, Beirut, North and South and El Qa’a) in Lebanon.

SM= *Serratia marcescens*, SA= *Salmonella* spp., AB= *Acinetobacter baumanii*, DE= *Delftia* spp., HA= *Hafnia*, KL= *Kluyvera ascorbata*, SH= *Shewanella* spp., PR= *Providencia* spp., RA= *Raoultella* spp., CI= *Citrobacter* spp., EB= *Enterobacter* spp., KP= *Klebsiella pneumoniae*, AE= *Aeromonas* spp., PA= *Pseudomonas* spp. EN= *Enterococcus* spp., EC= *E. coli*.

**Supplementary figure 2:** Pan-genome similarity, isolates data and maximum likelihood tree based on core genome SNPs of all sequenced *E. coli* isolates. Maximum likelihood phylogenetic tree is based on the core genome SNPs and was generated using FastTree 2.

Pan-genome is constructed using Roary based on the core and accessory genes showing phylogenetic relatedness between isolates, blue indicates present and white indicates absent fragment.

**Supplementary figure 3:** Pan-genome similarity, isolates data, maximum likelihood tree and core genome variation in recombination and SNPs for ST131 *E. coli* isolates. (a) Maximum likelihood phylogenetic tree based on core genome SNPs. (b) Recombination densities were detected between the samples using Gubbins.

**Supplementary figure 4:** Phages detected in whole genome sequenced isolates using PHASTER (phaster.ca/). Blue squares indicate the presence of the phage, orange square indicate that the phage carries *bla*_CTX-M-15_ and grey square indicates that the phage carries the increased serum resistance encoding gene *iss*. No phages were detected in isolates EC27, SM1 and PO1.

**Supplementary figure 5:** Distribution of the based on the source and time of collection.

## Notes

### Competing Interest Statement

The authors have declared no competing interest.

### Clinical Trial

Not a clinical trial

### Funding Statement

CNRS funding partially covered this work

