## Supplementary figures and images for "Detection of Antibiotic-Resistant Bacteria, Resistance Determinants, and Mobile Elements in Surface Waters in Lebanon"

### Supplemental Figure 1

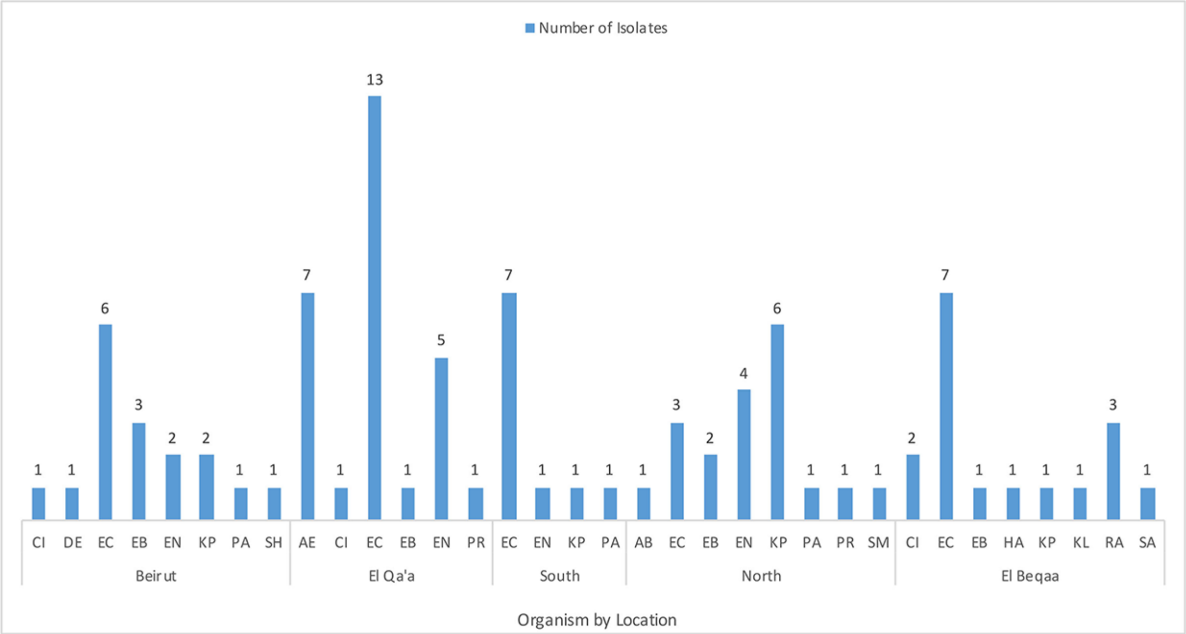

### Supplemental Figure 2

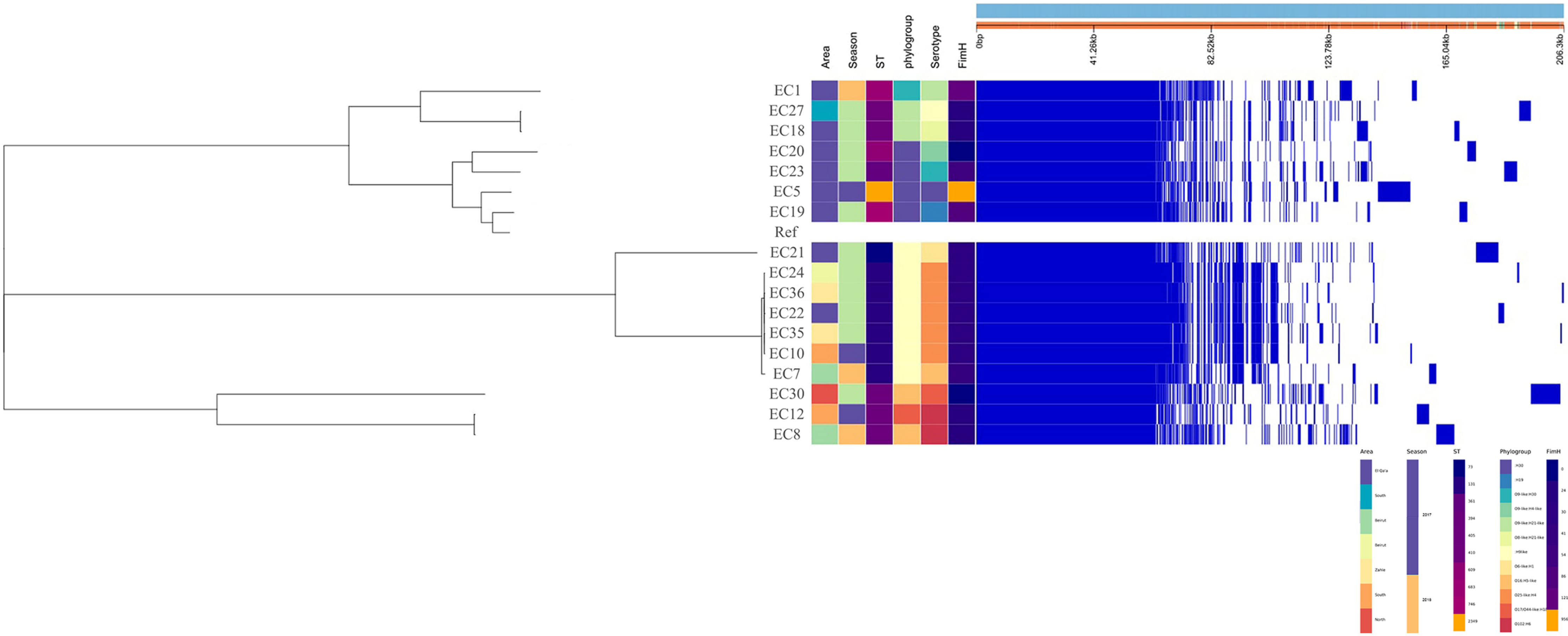

### Supplemental Figure 3

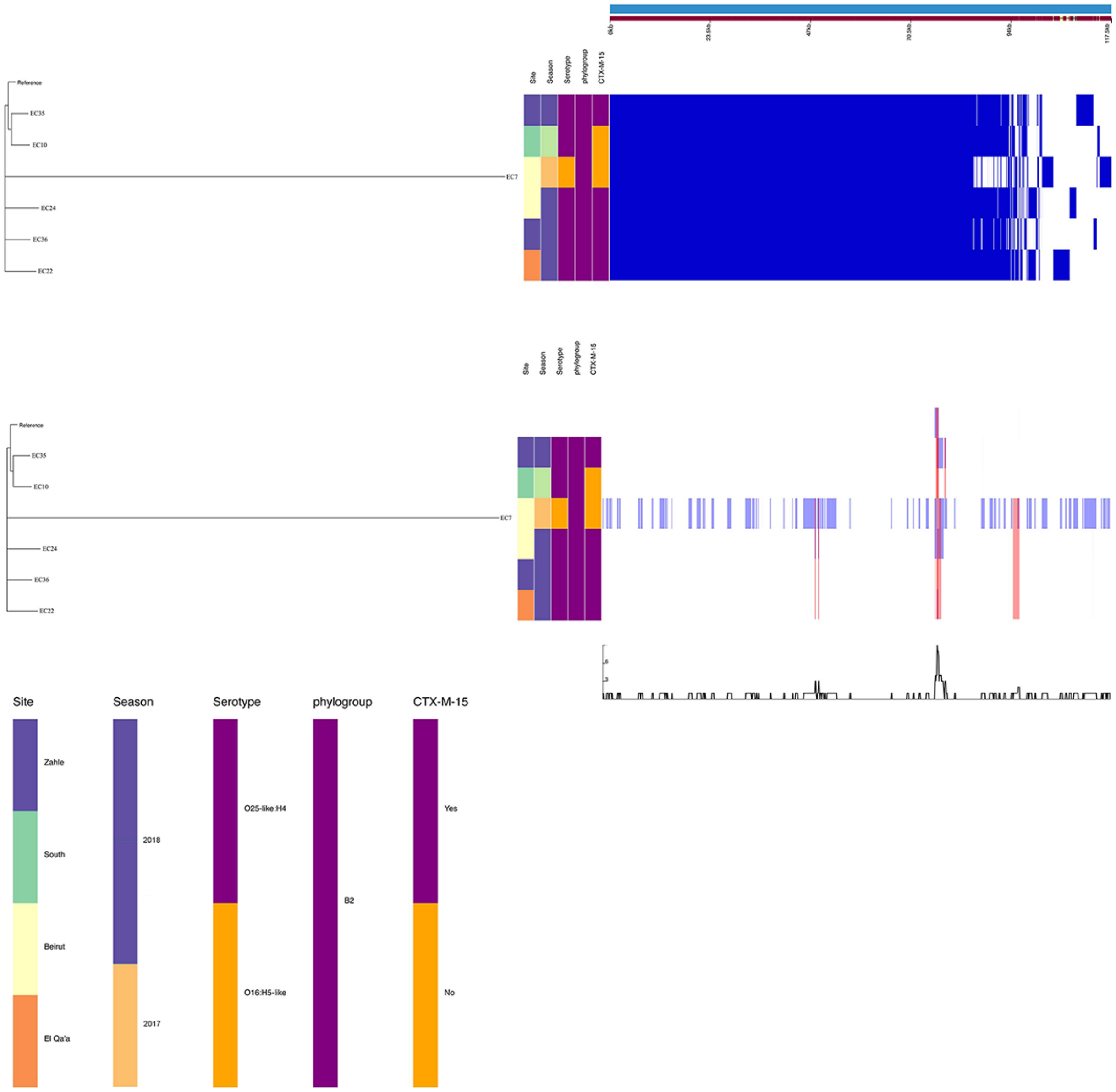

### Supplemental Figure 4

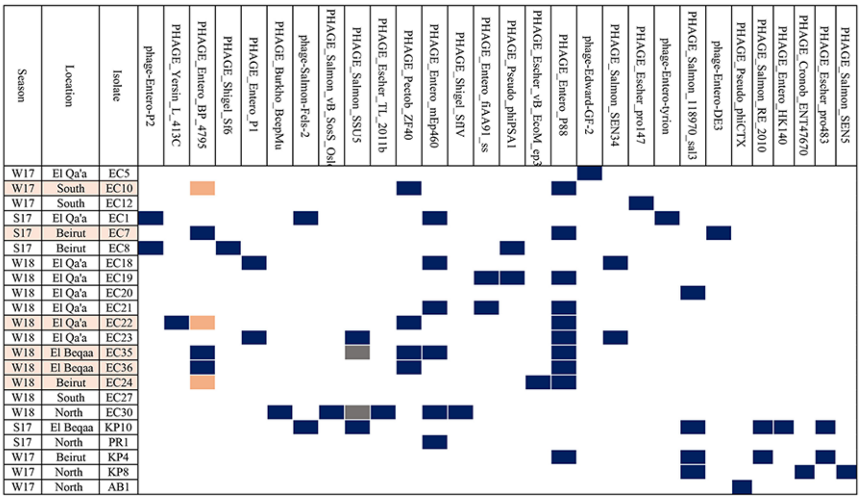
