## Supplemental Figure 5 for "Detection of Antibiotic-Resistant Bacteria, Resistance Determinants, and Mobile Elements in Surface Waters in Lebanon"

| Organism | Labeling | Season | Area | Organism | Labelings | Season | Area |
|---|---|---|---|---|---|---|---|
| *E. coli* | EC1 | 2017 | El Qa'a | *A. baumanii* | AB1 | 2017 | North |
| | EC2 - EC5 | 2017 | El Qa'a | *Aeromonas* spp. | AE1 - AE7 | 2017 | El Qa'a |
| | EC6 - EC8 | 2017 | Beirut | *Citrobacter freundii* | CI1 | 2017 | El Beqaa |
| | EC9 - EC12 | 2017 | South | | CI2 | 2017 | Al Qa'a |
| | EC13 - EC15 | 2017 | El Beqaa | | CI3 | 2017 | Beirut |
| | EC16 - EC23 | 2018 | El Qa'a | *Citrobacter gilleni* | CI4 | 2017 | El Beqaa |
| | EC24 - EC26 | 2018 | Beirut | Delftia spp. | DE1 | 2017 | Beirut |
| | EC27 - EC29 | 2018 | South | *Hafnia* | HA1 | 2017 | El Beqaa |
| | EC30 - EC32 | 2017 | North | *Kluyvera ascorbata* | KL1 | 2017 | El Beqaa |
| | EC33 - EC36 | 2018 | El Beqaa | *P. aeuroginosa* | PA1 | 2017 | Beirut |
| *K. pneumoniae* | KP1 - KP2 | 2017 | North | | PA2 | 2017 | North |
| | KP3 | 2017 | Beirut | *P. otitidis* | PO1 | 2017 | South |
| | KP4 | 2017 | Beirut | Providencia spp. | PR1 | 2017 | North |
| | KP5 | 2017 | South | | PR2 | 2017 | Al Qa'a |
| | KP6 - KP9 | 2017 | North | Raoultella spp. | RA1 - RA3 | 2017 | El Beqaa |
| | KP10 | 2017 | El Beqaa | *Salmonella enterica* | SA1 | 2017 | El Beqaa |
| Enterococcus spp. | EN1 | 2017 | El Qa'a | *Serratia macrescens* | SM1 | 2017 | North |
| | EN2- EN5 | 2017 | El Qa'a | Shewanella spp. | SH1 | 2017 | Beirut |
| | EN6 | 2017 | Beirut | *Enterobacter* spp. | EB1 | 2017 | Beirut |
| | EN7 | 2017 | Beirut | | EB2 - EB3 | 2017 | North |
| | EN8 | 2017 | South | | EB4 | 2017 | El Beqaa |
| | EN9 | 2017 | North | | EB5 | 2017 | El Qa'a |
| | EN10 - EN12 | 2017 | North | | EB6 - EB7 | 2017 | Beirut |
